# Nanoscale Podocyte Morphometrics Predict Disease Progression in IgA Nephropathy

**DOI:** 10.64898/2026.03.30.26349728

**Authors:** Robin Ebbestad, Arash Fatehi, Hannes Olauson, Katarzyna Bozek, Linus Butt, Thomas Benzing, Hans Blom, Hjalmar Brismar, Sigrid Lundberg, David Unnersjö-Jess

**Author notes:** SL & DUJ contributed equally.

## Abstract

**Introduction:** Podocyte injury is central to the pathogenesis of most glomerulonephritides (GN) and causes segmental glomerulosclerotic lesions that predict progression in IgA Nephropathy (IgAN). Recent advances in high-resolution microscopy and AI-assisted image analysis have enabled detailed quantification of podocyte foot process (FP) morphology. However, whether nanoscale podocyte morphometrics can predict disease progression or treatment response in GN has not been investigated.

**Aim:** To evaluate whether nanoscale podocyte morphometric parameters predict clinical characteristics, disease progression, and treatment response in GN, with a focus on IgAN.

**Method:** Podocyte morphometrics were analyzed in kidney biopsies from patients with GN using high-resolution microscopy and the deep learning-based tool Automatic Morphometric Analysis of Podocytes (AMAP). Four morphometric parameters were quantified: slit diaphragm length (SDL), FP area, FP circularity and FP perimeter. These parameters were correlated with clinical characteristics, conventional electron microscopy (EM) findings and longitudinal follow-up data.

**Results:** The study included 37 patients with GN from Danderyd University Hospital (Stockholm, Sweden), with IgAN representing the largest diagnostic subgroup (n = 19). The median follow-up for the cohort was 3.0 years. SDL correlated significantly with urine albumin-to-creatinine ratio (uACR; p = 0.021), whereas conventional EM measurements did not (p = 0.22). Within the IgAN subgroup, lower SDL was associated with a steeper decline in eGFR, higher FP area with increased long-term proteinuria, and higher FP circularity with improvement in uACR during the first year. The association between lower SDL and eGFR decline remained as a trend in IgAN patients not treated with corticosteroids (p = 0.068) but was absent in the treatment group (p = 0.59).

**Conclusion:** In this proof-of-concept study, nanoscale podocyte morphometrics demonstrated greater sensitivity than conventional EM in quantifying podocyte injury and predicting progression in IgAN. These findings suggest that high-resolution morphometrics may improve risk stratification in IgAN but require validation in larger, independent cohorts before clinical implementation.

## Introduction

Kidney biopsy remains the gold standard diagnostic evaluation of suspected glomerular disease.^1^ Beyond the primary pathology diagnosis, semi-quantitative histological scoring systems are used to assess patterns of injury. While these scores provide prognostic information supplementary to clinical data, manual scoring is limited by significant time requirements and interobserver variability.^2–7^ To overcome these challenges, several groups have developed deep learning-based algorithms to automate the classification of digitized histology slides and have identified novel morphological biomarkers.^8–11^ However, standard histology techniques are inherently restricted in resolution and molecular information, which limits the depth of data that can be extracted.

In this work we focus on the clinical application of nanoscale morphometric analysis of podocyte injury. The podocyte is a principal cell type of the glomerulus that maintains the structural integrity of the filtration barrier. According to current models, podocytes generate buttressing forces that compress the glomerular basement membrane (GBM), and disruption of this mechanical function leads to proteinuria.^12,13^ While central to the development of disease phenotypes such as focal segmental glomerulosclerosis (FSGS), podocyte injury is present in most glomerular diseases. Recently, it has been shown that histological subclasses of FSGS lesions in IgA Nephropathy (IgAN) predict both disease progression and response to corticosteroid treatment, indicating that podocyte injury should be further studied in glomerular diseases not primarily considered as podocytopathies.^14,15^

In recent years, super-resolution immunofluorescence microscopy has enabled the visualization and quantification of podocyte foot process (FP) morphology, opening a new field of nanoscale podocyte morphometrics in kidney disease.^16–20^ In 2023, our group developed a deep learning-based approach to analyze podocyte morphometry in Minimal Change nephropathy (MCD), establishing the methodological foundation for quantitative FP analysis. Using this approach, we identified distinct combinations of morphometric parameters, such as slit diaphragm length (SDL), FP area and FP circularity, that were associated with response to different treatments.^21^ In a complementary approach, Siegerist et al have also demonstrated differences in slit density distributions between primary and secondary FSGS.^22^ However, whether nanoscale podocyte morphometrics can predict clinical outcomes longitudinally in GN, particularly in diseases not traditionally classified as podocytopathies, has not been investigated.

In the present study, we applied high-resolution confocal microscopy and the deep learning-based tool Automatic Morphological Analysis of Podocytes (AMAP)²¹ to kidney biopsies from patients with IgAN, Membranous Nephropathy (MN) and Lupus Nephritis (LN), also including patients with thin basement membrane disease (TBMD) and nephrosclerosis (HTN) as controls. We demonstrate that nanoscale podocyte morphometrics, including slit diaphragm length (SDL), FP area and FP circularity, identify associations with disease progression and treatment response that are not captured by conventional histology or electron microscopy (EM).

## Material & Methods

For detailed information about patient selection, sample preparation and image analysis, see Supplementary information.

The present study is a retrospective analysis of kidney biopsy samples from 37 patients who had been included in the BIONEF-DS patient cohort at the Nephrology clinic, Danderyd University Hospital, Sweden. This cohort consists of patients who at the time of their clinical indicated biopsy had consented to the use of left-over renal tissue, extra blood and urine samples and prospectively collected clinical data for renal histology and biomarker research. Information on creatinine, proteinuria, comorbidities, medication and other common clinical parameters were collected at baseline. Follow-up data was collected through electronic medical records. All decisions on the frequency of follow-up visits, laboratory tests and therapy were done at the discretion of the treating physician. The event of end stage kidney disease (ESKD) was recorded, defined as eGFR below 15 mL/min/1.73 m² or start of kidney replacement therapy (KRT) by dialysis or transplantation. Patients were followed until September 2025, start of KRT or loss to follow-up.

OCT-frozen biopsy specimens were fixed in PFA, cleared in 4% SDS w/ boric acid and fluorescently immunolabelled for nephrin (R&D Systems, cat. no. AF4269) to visualize the slit diaphragm and podocyte foot processes (FPs) based on the protocol described by Unnersjö-Jess et al (details available in Supplement Methods).^17^ High-resolution confocal image stacks of the immunolabelled biopsy sections were acquired using a Leica SP8 confocal microscope (Leica Microsystems, Wetzlar, Germany). The image stacks were maximum intensity projected before morphometric image analysis.

To enhance segmentation results, images with low absolute signal intensities were subjected to pre-processing using ImageJ/Fiji:s^23^ auto adjust brightness/contrast function or local contrast enhancement (CLAHE), depending on local variation. Images with low signal to background ratio were subjected to deconvolution in Huygens software using either Classic Maximum Likelihood Estimation or Classical Tikhonov-Miller deconvolution (Scientific Volume Imaging, The Netherlands, http://svi.nl).

To quantify podocyte morphometrics, the previously published AMAP model was used.^21^ This model features a modified U-Net architecture with a combined discriminative loss and cross-entropy loss function which segments both the slit diaphragm and each individual foot process (FP) for morphometric analysis.^21,24^ The original model was fine-tuned on new images from this study before the final analysis. Further details on the fine-tuning process are available in the Supplementary Methods. The final segmentation results were post-processed by removing all FPs <0.1µm^2^ and filling in holes in automatic image segmentations.

Four distinct morphometric parameters were extracted via automatic segmentation. Slit diaphragm length (SDL) was quantified to assess slit coverage, while FP area, perimeter and circularity were calculated to characterize individual FP morphology. SDL was defined as the total length of the slit diaphragm normalized to the imaged capillary surface area. This reference area was automatically assigned as described in the AMAP protocol.^21^

FP circularity was expressed as a value between 0 to 1, where 1 represents a perfect circle and values approaching 0 indicate an increasingly elongated morphology. FP area was defined as the measured surface area of each individual FP, while FP perimeter represents the total boundary length. Due to the strong co-linearity between area and perimeter, the latter was excluded from most analyses. Morphometric data were recorded both at the individual FP level and as a global mean per patient. Unless otherwise stated, the patient-level mean was used for correlative analyses.

The longitudinal eGFR slope was chosen as the main outcome and was estimated for all patients using linear regression with the ordinary least squares method. To maintain statistical reliability, eGFR slope was only estimated for patients with a minimum of three eGFR data points.

All information on proteinuria was obtained by urine albumin-to-creatinine ratio (uACR) measurements. Time-average uACR (TA uACR) was calculated with log-transformed values using trapezoidal geometric mean averaging between follow-up visits. Any patient with less than 3 measurements of uACR was excluded from TA uACR estimation.

The 1-year %-change in uACR was calculated between the mean baseline uACR at 0-30 days and the mean uACR at 12 months (±3 months). Since follow-up visit timing were not regularized, this windowed mean approach was employed to maximize patient capture, reduce the effect of random fluctuations and avoid selection bias.

In the largest diagnosis group, IgAN, morphometric features were analyzed for correlations with eGFR slope, uACR, TA uACR and %-change in uACR during the first year of follow-up. Subgroup analysis was performed concerning eGFR slope and %-change in uACR, stratified by if the patient received treatment with corticosteroids (CS) or not.

## Results

### Clinical characteristics of the study cohort

37 patients were included in the study and had a diagnosis of IgA nephropathy (IgAN, n = 19), IgA vasculitis with nephritis (IgAVN, n = 3), membranous nephropathy (MN, n = 7), lupus nephritis (LN, n = 3), thin basement membrane disease (TBMD, n = 2, none with confirmed genetic alteration) and nephrosclerosis (HTN, n = 3). The median follow-up for the cohort was 3.0 years. Two patients reached ESKD during follow-up, both with IgAN, and one additional patient, with MN, lost more than 50% in eGFR (Table 1).

**Table 1.** Baseline characteristics, treatment at biopsy, eGFR slope, time-averaged uACR and podocyte morphometrics for the whole cohort. Continuous variables are presented as either median with interquartile range (IQR) or mean ± standard deviation (SD). For variables where there are missing values for a subset of patients, the number of patients with a valid measurement is stated in parentheses in the table. *AU = Arbitrary units*.

|  | Total<br>(n = 37) | IgAN<br>(n = 19) | IgAV<br>(n = 3) | MN<br>(n = 7) | LN<br>(n = 3) | TBMD<br>(n = 2) | HTN<br>(n = 3) |
| --- | --- | --- | --- | --- | --- | --- | --- |
| **Baseline characteristics** |  |  |  |  |  |  |  |
| **Age, years (median [IQR])** | 44.0 (36.0-59.0) | 43.0 (35.0-50.5) | 39.0 (36.5-47.5) | 61.0 (47.5-71.5) | 32.0 (31.0-36.5) | 40.0 (35.5-44.5) | 70.0 (64.5-70.5) |
| **Sex (Male:Female, %)** | 23:14, 62.2% | 15:4, 78.9% | 2:1, 66.7% | 5:2, 71.4% | 0:3, 0.0% | 0:2, 0.0% | 1:2, 33.3% |
| **eGFR at biopsy, mL/min/1.73m<sup>2</sup> (mean ± SD)** | 72.9 ± 26.0 | 64.4 ± 26.7 | 78.1 ± 12.2 | 85.8 ± 24.1 | 82.0 ± 22.3 | 106.3 ± 10.6 | 59.9 ± 21.4 |
| **uACR at biopsy, mg/mmol (median [IQR])** | 66.7 [38.1–174.6] | 67.6 [44.9–101.5] | 39.6 [37.4–43.7] | 243.2 [114.9–357.8] | 38.1 [28.0–163.8] | 82.4 [42.2–122.5] | 1.7 [1.1–101.3] |
| **MAP at biopsy, mmHg (mean ± SD)** | 98.0 ± 9.0 | 97.4 ± 8.1 | 100.8 ± 5.6 | 99.2 ± 7.8 | 91.4 ± 15.8 | 94.0 ± 15.1 | 105.8 ± 10.5 |
| **BMI, kg/m<sup>2</sup> (median [IQR])** | 24.6 (23.6-28.7)<br>(n = 33) | 24.3 (23.5-28.7) | 28.3 (27.5-29.1)<br>(n = 2) | 24.6 (22.8-27.6) | 23.6 (NA)<br>(n = 1) | 25.6 (NA)<br>(n = 1) | 26.8 (25.3-28.8) |
| **Hypertension, %** | 62.2 | 73.7 | 0.0 | 71.4 | 0.0 | 50.0 | 100.0 |
| **Follow-up & outcome** |  |  |  |  |  |  |  |
| **Follow-up, years (median [IQR])** | 3.0 [2.4–3.8] | 3.3 [2.6–3.5] | 2.1 [1.4–3.3] | 2.8 [2.3–4.4] | 2.4 [1.2–3.7] | 3.8 [3.2–4.3] | 3.0 [1.5–3.5] |
| **Yearly eGFR slope (mean ± SD)** | -2.8 ± 7.8<br>(n = 32) | -3.9 ± 7.3<br>(n = 17) | 7.0 ± 8.4 | -5.2 ± 9.4 | -2.6 ± NA<br>(n = 1) | -2.7 ± 2.1 | 0.2 ± 0.2<br>(n = 2) |
| **Time-averaged uACR, mg/mmol (median [IQR])** | 32.4 [16.1–62.8]<br>(n = 31) | 44.9 [17.6–63.0]<br>(n = 16) | 22.5 [18.4–29.4] | 20.8 [17.8–117.6] | 32.4 [32.4–32.4]<br>(n = 1) | 34.6 [17.9–51.3] | 10.7 [5.6–15.7]<br>(n = 2) |
| **ESKD, %** | 5.4 | 10.5 | 0.0 | 0.0 | 0.0 | 0.0 | 0.0 |
| **Treatment at baseline** |  |  |  |  |  |  |  |
| **RAAS inhibition (ACEi or ARB), %** | 59.5 | 73.7 | 0.0 | 71.4 | 33.3 | 50.0 | 33.3 |
| **SGLT2i, %** | 8.1 | 15.8 | 0.0 | 0.0 | 0.0 | 0.0 | 0.0 |
| **Immunosuppression, %** | 10.8 | 0.0 | 5.3 | 0.0 | 0.0 | 100.0 | 0.0 |
| **Podocyte morphometrics** |  |  |  |  |  |  |  |
| **SDL, μm/μm<sup>2</sup> (mean ± SD)** | 1.13 ± 0.24 | 1.24 ± 0.19 | 1.32 ± 0.05 | 0.82 ± 0.14 | 1.02 ± 0.18 | 1.10 ± 0.26 | 1.17 ± 0.19 |
| **FP area, μm<sup>2</sup> (mean ± SD)** | 0.26 ± 0.03 | 0.24 ± 0.03 | 0.24 ± 0.01 | 0.30 ± 0.02 | 0.28 ± 0.01 | 0.30 ± 0.03 | 0.22 ± 0.00 |
| **FP circularity, AU (mean ± SD)** | 0.53 ± 0.03 | 0.52 ± 0.02 | 0.51 ± 0.03 | 0.56 ± 0.01 | 0.53 ± 0.04 | 0.54 ± 0.04 | 0.50 ± 0.02 |

The mean yearly eGFR slope for the total study population was −2.8 ml/min/year with the most negative mean eGFR slope in the MN group (−5.2 ml/min/y). The median uACR at biopsy was 66.7 mg/mmol (590 mg/g), highest in the MN group (243.2 mg/mmol [2150 mg/g]). During follow-up, uACR decreased overall and the median TA uACR across the cohort was 32.4 mg/mmol (286 mg/g).

At the time of biopsy, 59.5% of the patients were on renin-angiotensine system blockade (RASB), and 3 patients (all with IgAN) were treated with an SGLT2-inhibitor (SGLT2i). One of the IgAN patients and all 3 LN patients were on immunosuppressive treatment (Table 1).

### Overview of the podocyte morphometric readouts

Figure 1 shows an overview of the workflow (Figure 1a) with examples of the segmentation results (Figure 1b-d) and schematic images of the morphometric readouts (Figure 1e-g). In total, 439 images were analyzed for morphometric features with a median number of 463 segmented foot processes per patient. The mean number of analyzed glomeruli was 3.0 per patient for the whole cohort and 2.4 per patient for IgAN.

**Figure 1.**
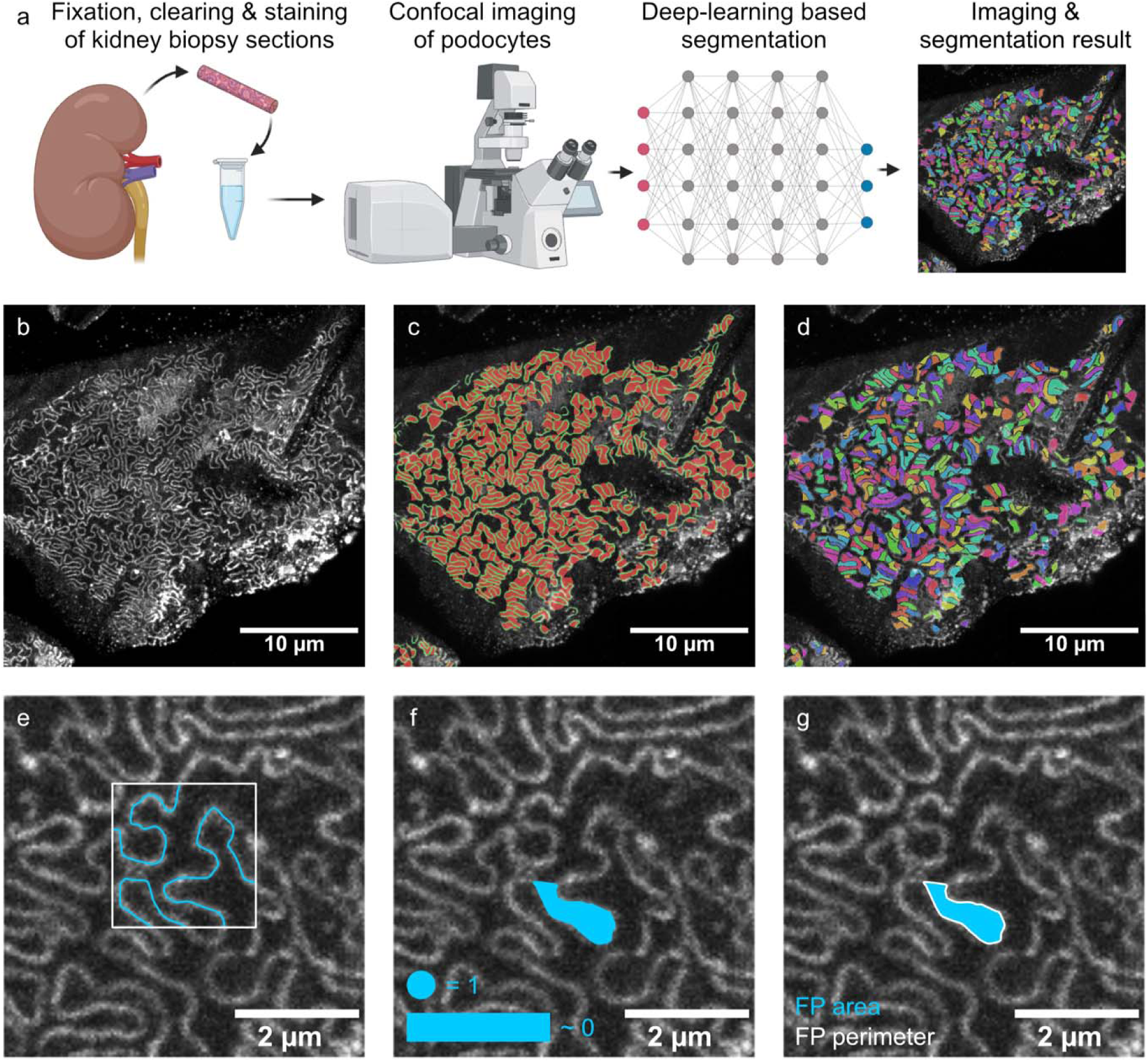
Overview of the analysis workflow and the morphometric parameters. (a) OCT frozen kidney biopsies were sectioned, cleared and immunolabelled followed by confocal imaging and deep-learning segmentation. (b) Maximum intensity projection confocal image of nephrin visualizing the slit diaphragm network and foot processes. (c) Output of the semantic segmentation of both the slit diaphragm and foot processes. Foot processes are marked in red and the slit diaphragm in green. (d) Output of the instance segmentation resulting in segmentation of each individual foot process in the image. Each foot process is presented in an individual color. (e-g) From the 2 segmentation outputs, the morphometric parameters SDL (e), FP circularity (f), FP area and FP perimeter (g) are extracted. (e) Schematic image of how SDL is measured. The length of the slit (in blue) is divided by the surface area (outlined in white) resulting in a length-density measurement of slit coverage. (f) Schematic image of how FP circularity is measured. From the segmentation of the individual foot processes, the FP circularity is calculated using the formula, circularity = 4 π (area/perimeter−2). The result is a value between 0 and 1 where a perfect circle has a circularity of 1 and a very elongated rectangle approaches 0. (g) Foot process size is calculated both by FP area and by FP perimeter. The FP area is the pixel area of each segmented foot process converted to µm^2^ and the FP perimeter is the outline of the foot process with it closed at the base.

In the analyzed images, the features SDL, FP area and FP circularity captured visually different alternations in podocyte architecture (Figure 2). Progressively lower SDL showed decreasing capillary surface coverage of the slit diaphragm, mirroring effacement (Figure 2a-c). For FP area, images of median FP area showed the visually most healthy FPs while low FP area and high FP area showed signs of podocyte injury (Figure 2d-f). FP circularity showed a pattern ranging from thin elongated FPs for low FP circularity to short and wide FPs for high FP circularity (Figure 2g-i).

**Figure 2.**
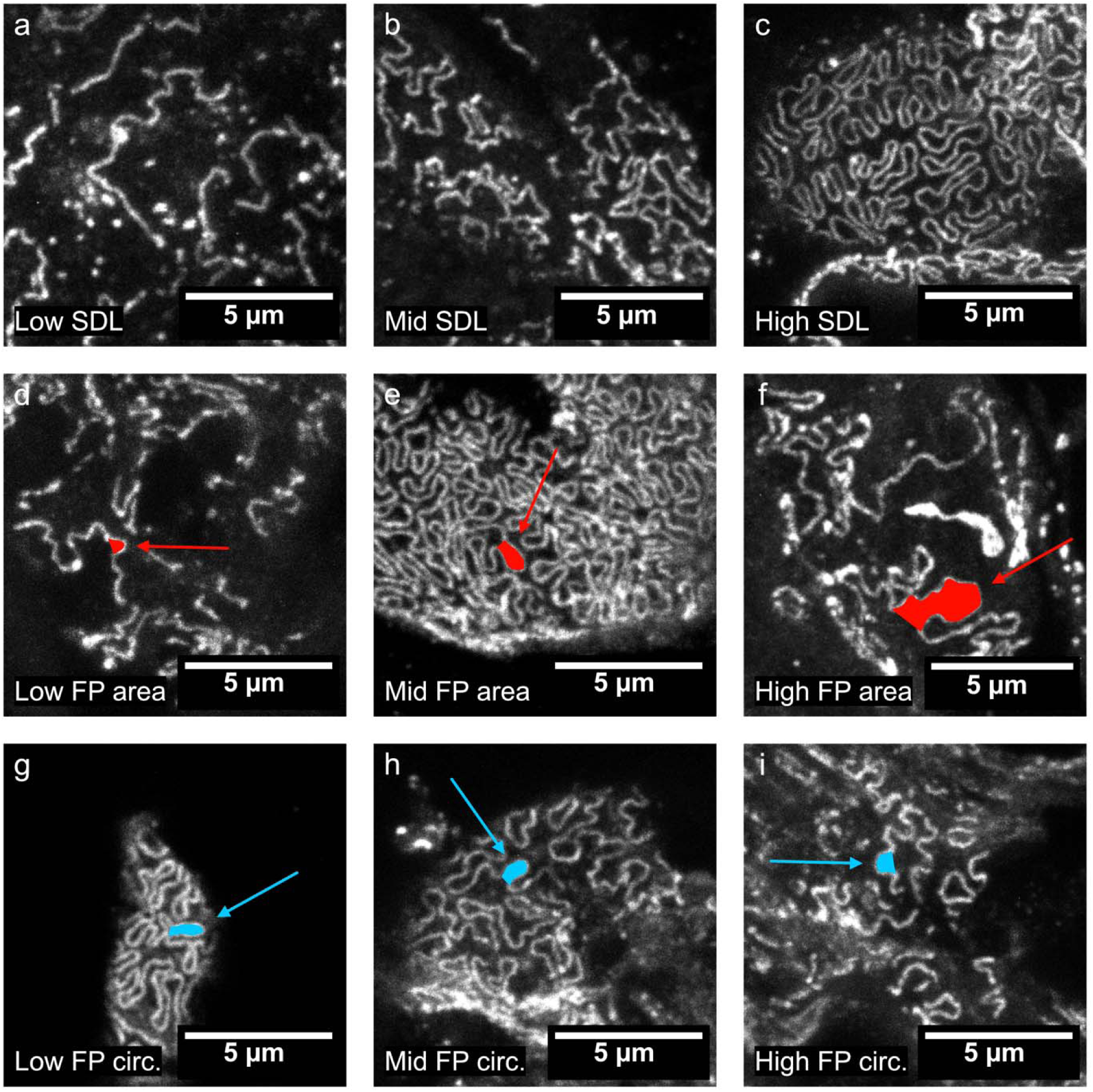
Different podocyte morphometric features capture different alterations in podocyte architecture. The images in the figure show example images which have mean values that are among the lowest, median and highest for each podocyte morphometric feature. (a, b, c) SDL closely resembles FP effacement. Low SDL (a) shows severe effacement with little organization or interdigitation of podocyte FPs. With increasing SDL (b, c) there is an increasing degree of organization. (d, e, f) FP area shows a different distribution with the visually most healthy FPs in the median example (e). Low FP area (d) and high FP area (f) both displays altered FPs that are either abnormally small or large. (g, h, i) FP circularity ranges from thin elongated FPs with low FP circularity (g) to short and wide FPs with high FP circularity (i). The median FP circularity (h) image displays a variation of FP shapes that are largely somewhere between the low (g) and high (i) FP circularity example.

### Correlation of podocyte morphometrics to baseline clinical parameters, histology findings and treatment in the whole study cohort

#### Correlation to clinical baseline characteristics

In the whole cohort, there was a non-significant trend for lower SDL correlating with higher baseline uACR, a significant correlation between FP circularity and baseline uACR but no correlation between FP area and baseline uACR (Table 2). Higher FP area had a strong correlation with higher baseline eGFR (p = 0.006) but there was no correlation between eGFR and SDL or FP circularity (Table 2). All other clinical parameters that were tested had no correlation with the morphometric parameters.

**Table 2.** Correlations between podocyte morphometric features and clinical characteristics, histological parameters and treatment at baseline. Unless otherwise stated in parentheses next to the variable name, the variables are tested for correlations in the whole cohort. Shapiro-Wilk test was used as normality test for all variables. For normally distributed variables, a parametric test of either Pearson (continous variables) or T-test (non-continous variables) was performed. For non-normal variables, a non-parametric test of Spearman Rank (continuous variables) or Mann-Whitney U-test (non-continous variables) was performed. All results from non-parametric test are marked with “†”. Statistically significant correlations are marked with “*” for p-values < 0.05 and “**” for p-values < 0.01. *AU = Arbitrary units*.

| | SDL, $\mu\text{m}/\mu\text{m}^2$ | FP area, $\mu\text{m}^2$ | FP circularity, AU |
| --- | --- | --- | --- |
| **Clinical Parameters** |  |  |  |
| Age, years | -0.280†<br>(p=0.093) | 0.121†<br>(p=0.475) | 0.287† (p=0.085) |
| Sex, Male:Female | +0.029<br>(p=0.709) | -0.013 (p=0.257) | +0.008 (p=0.402) |
| eGFR, mL/min/1.73m<sup>2</sup> | -0.190 (p=0.261) | 0.442**<br>(p=0.006) | 0.003 (p=0.987) |
| MAP, mmHg | 0.037 (p=0.828) | 0.063 (p=0.712) | -0.147 (p=0.387) |
| BMI, kg/m<sup>2</sup> | 0.080†<br>(p=0.660) | -0.062†<br>(p=0.732) | -0.066† (p=0.714) |
| uACR, mg/mmol | -0.315†<br>(p=0.058) | 0.219†<br>(p=0.193) | 0.351*† (p=0.033) |
| Hypertension, Yes:No | -0.045 (p=0.598) | -0.001 (p=0.914) | +0.001 (p=0.928) |
| **Histological Parameters** |  |  |  |
| GSG% | 0.135†<br>(p=0.425) | -0.444**†<br>(p=0.006) | -0.063† (p=0.712) |
| GSG% (IgAN only) | 0.041† (p=0.867) | -0.620**†<br>(p=0.005) | 0.212† (p=0.385) |
| Oxford M (IgAN only) | +0.095<br>(p=0.159) | -0.010†<br>(p=0.491) | +0.015* (p=0.027) |
| Oxford E (IgAN only) | +0.038<br>(p=0.680) | +0.001†<br>(p=0.904) | +0.003 (p=0.717) |
| Oxford S (IgAN only) | +0.085<br>(p=0.303) | -0.003†<br>(p=0.211) | -0.009 (p=0.342) |
| Oxford T (IgAN only) | 0.228†<br>(p=0.347) | -0.350†<br>(p=0.142) | 0.379† (p=0.110) |
| Oxford C (IgAN only) | 0.109†<br>(p=0.657) | 0.153†<br>(p=0.532) | 0.022† (p=0.929) |
| **Treatment at baseline** |  |  |  |
| RAASi (ACEi/ARB), Yes:No | -0.081 (p=0.333) | +0.012<br>(p=0.257) | +0.002 (p=0.821) |
| SGLT2i, Yes:No | +0.055<br>(p=0.336) | -0.001 (p=0.958) | -0.012* (p=0.020) |
| Immunosuppression, Yes:No | -0.053 (p=0.641) | +0.011<br>(p=0.328) | +0.001 (p=0.949) |

#### Correlation to routine histology findings and electron microscopy

The percentage of globally sclerosed glomeruli (GSG%) has been shown to correlate closely with eGFR and to be predictive of outcome in glomerular disease.^25^ Higher FP area had a strong correlation with a lower GSG% in both the whole cohort and in IgAN (Table 2). This was in line with the correlation between higher FP area and higher baseline eGFR. SDL and FP circularity had no correlation with GSG%. In IgAN, the only correlation between MEST-C scores^3^ and morphometric features were a positive correlation between M-score and FP circularity (Table 2).

Biopsies from 16 patients had been examined by electron microscopy (EM) as part of the clinical evaluation. This subgroup included all diagnoses enrolled in the study [MN (n = 6), IgAN (n = 3), HTN (n = 2), LN (n = 2), TBMD (n = 2) and IgAV (n = 1)]. Based on the written biopsy reports, patients were categorized into three groups depending on the degree of podocyte foot process effacement: no to mild/focal, moderate or severe effacement. 2 patients had varying effacement between two groups and were sorted based on the highest severity in the report.

There was no significant difference in uACR between the three effacement groups (Figure 3a & 3b) despite a tendency to higher mean uACR with moderate or severe effacement. Lower SDL however correlated significantly with higher uACR in the same group with available EM reports. (Figure 3c). Comparing the degree of effacement with the morphometrics features showed that the group with severe effacement had significantly lower SDL and higher FP circularity than the group with no to mild/focal effacement (Figure 3d-f).

**Figure 3.**
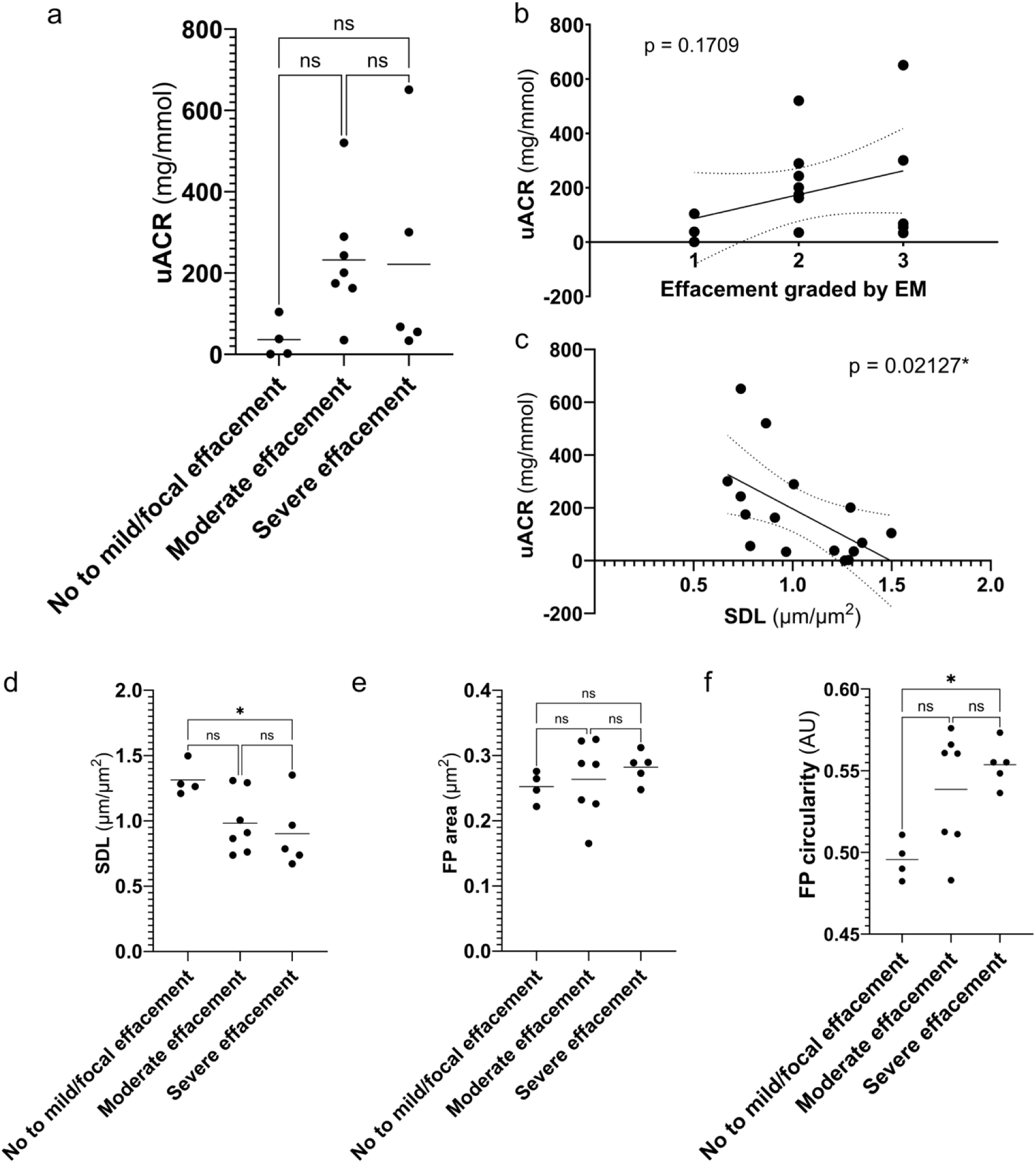
Effacement graded by electron microscopy compared to assessment by podocyte morphometrics and correlation to baseline uACR. All 16 patients whose biopsies had undergone clinical evaluation by electron microscopy were grouped into No to mild/focal, Moderate and Severe effacement groups based on the biopsy report from the clinical pathologist. (a) Effacement groups were tested for correlation with baseline uACR using one-way ANOVA, followed by Tukey’s multiple comparisons test. No significant differences in baseline uACR could be found between the effacement groups (p = 0.2183), but there was a non-significant trend for higher mean uACR with Moderate and Severe effacement. (b) The effacement groups were also converted to an ordinal scoring from 1 to 3, from lowest to highest degree of effacement, and tested for correlation to baseline uACR by linear regression analysis. Similarly, there was no significant correlation between effacement grade and uACR by linear regression analysis as well. (c) In contrast, lower SDL was associated with higher baseline uACR, in the same subgroup, by linear regression analysis. (d, e, f) Effacement, graded by electron microscopy, was also compared to the morphometric parameters directly by one-way ANOVA, followed by Tukey’s multiple comparisons test. (d) SDL was significantly lower (p = 0.046) and (f) FP circularity was significantly higher (p = 0.014) in the group with the most severe effacement compared to the least severe. There was no correlation between effacement grade and FP area (e). *AU = Arbitrary units*.

#### Correlation to treatment at baseline and decision to treat with corticosteroids

In the whole cohort, patients treated with SGLT2i had lower FP circularity but there were otherwise no significant correlations between baseline treatment with RASB, SGLT2i or Immunosuppression and morphometric features (Table 2).

In IgAN, none of the new morphometric features was associated with the decision to treat with CS during follow-up. The only baseline factor associated with CS-therapy was a positive C-score (Suppl. Table 1).

### Distribution and Cross-correlation of morphometric features depending on diagnosis

The morphometric features showed varying degrees of interdependence, with certain correlations present across the entire cohort while others were specific to diagnosis subgroups. In the total cohort, a lower SDL correlated with both higher FP area and higher FP circularity (Suppl. Figure 1a & 1b). While no significant correlation was found between FP area and FP circularity in the whole cohort, a significant inverse correlation was found for the IgAN subgroup (Suppl. Figure 1c).

Exploring patient clustering using principal component analysis of all morphometric features revealed clustering of all MN patients with one patient with LN (classified as WHO 5, i.e., membranous form of LN) and one with TBMD (with 162.7 mg/mmol uACR at biopsy increasing to 346.5 mg/mmol by the last follow-up) (Suppl. Figure 1d). This indicates that the podocyte morphometrics identifies patients with significant podocytopathy. By contrast, the TBMD patient in the cluster was described as only having mild to moderate FP effacement in the EM report.

The distribution of FP area, for all individual FPs, varied significantly between most diagnoses (Suppl. Figure 2). The most pronounced differences were seen between MN and the diagnoses; HTN, IgAN and IgAVN (Suppl. Figure 2a-b). Differences in FP area were primarily driven by a rightward shift in the distribution tail, reflecting an increased frequency of enlarged FPs in diagnoses such as MN. However, the peak of the FP area distribution (the mode), remained similar across most diagnoses (Suppl. Table 2.). The distribution of FP circularity was also significantly different between most diagnoses with MN having the highest mean FP circularity (Suppl. Figure 2c-d).

### Morphometric features in IgAN patients in correlation to follow-up data and CS treatment

#### The IgAN cohort

Two patients in the IgAN group were lost to follow-up before 3 eGFR-measurements had been obtained and were thus excluded from the primary outcome analysis (eGFR slope). Median follow-up for the remaining 17 patients was 3.3 years and the mean eGFR slope was −3.94 mL/min/1.73m2/year. 7 patients were treated with CS during follow-up. For uACR analyses, at baseline or during follow-up, one patient was excluded due to having less than 3 uACR measurements.

#### SDL predicts eGFR slope in IgAN

Lower SDL correlated significantly with a more negative eGFR slope in IgAN (Figure 4a). In subgroups stratified by CS therapy, the same trend remained in the untreated subgroup but did not reach statistical significance (Figure 4b). No association between SDL and eGFR slope was seen in the CS treated group (Figure 4c). SDL was not significantly correlated with baseline uACR, 1-year %-change in uACR or TA uACR in IgAN (Suppl. Figure 3).

**Figure 4.**
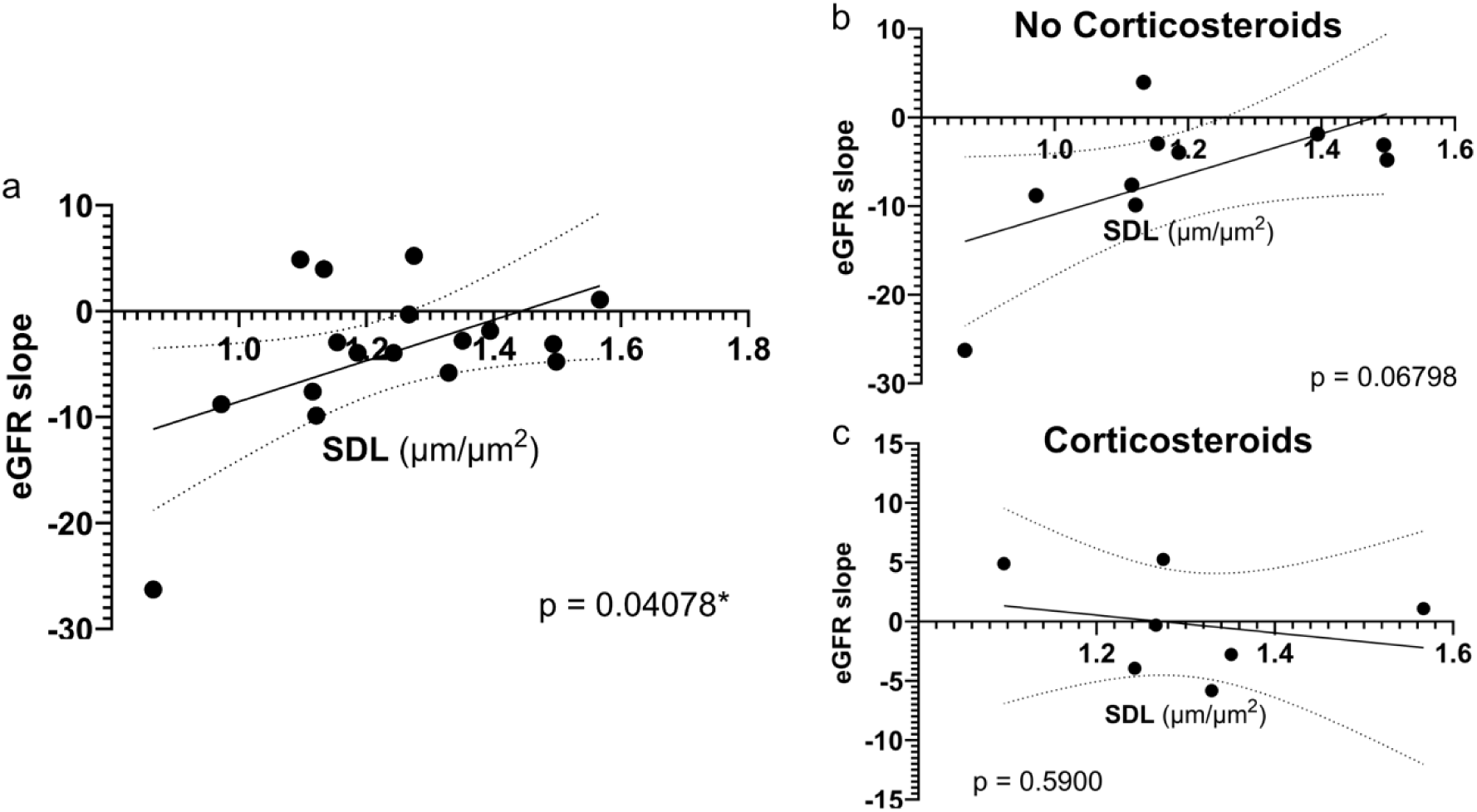
Correlation between SDL and eGFR slope in IgA Nephropathy, including subgroup analysis stratified by if the patient received corticosteroid therapy. The graphs show scatterplots with a fitted linear regression for the relationship between SDL and eGFR slope. Each point re++presents the mean value for one patient. eGFR slope was measured as the yearly change in eGFR (mL/min/1.73m^2^/year). Correlations were tested by linear regression analysis. (a) Lower SDL was significantly correlated with a more negative eGFR slope. (b) A trend remained for lower SDL being associated with worse eGFR slope in untreated IgAN-patients, but this did not reach statistical significance. (c) There was no correlation between SDL and eGFR slope among IgAN-patients who received corticosteroids.

#### FP area correlates with both eGFR slope and TA uACR in IgAN

FP area exhibited a non-linear, inverse U-shaped relationship with eGFR slope, where both low and high FP area values were associated with more rapid eGFR decline (Figure 5). This parabolic trend was confirmed by a quadratic regression model, which showed a significantly better fit than linear regression even after a sensitivity analysis excluding the most negative outlier (Figure 5a-b). However, no correlation was found between FP area and eGFR slope within CS subgroups. FP area did not correlate with baseline uACR or 1-year %-change in uACR but was strongly positively correlated with TA uACR (Figure 5c-d). In CS subgroups, there was a non-significant trend towards higher FP area being associated with worsening uACR during the 1st year, a pattern absent in the subgroup receiving CS therapy (Figure 5e-f).

**Figure 5.**
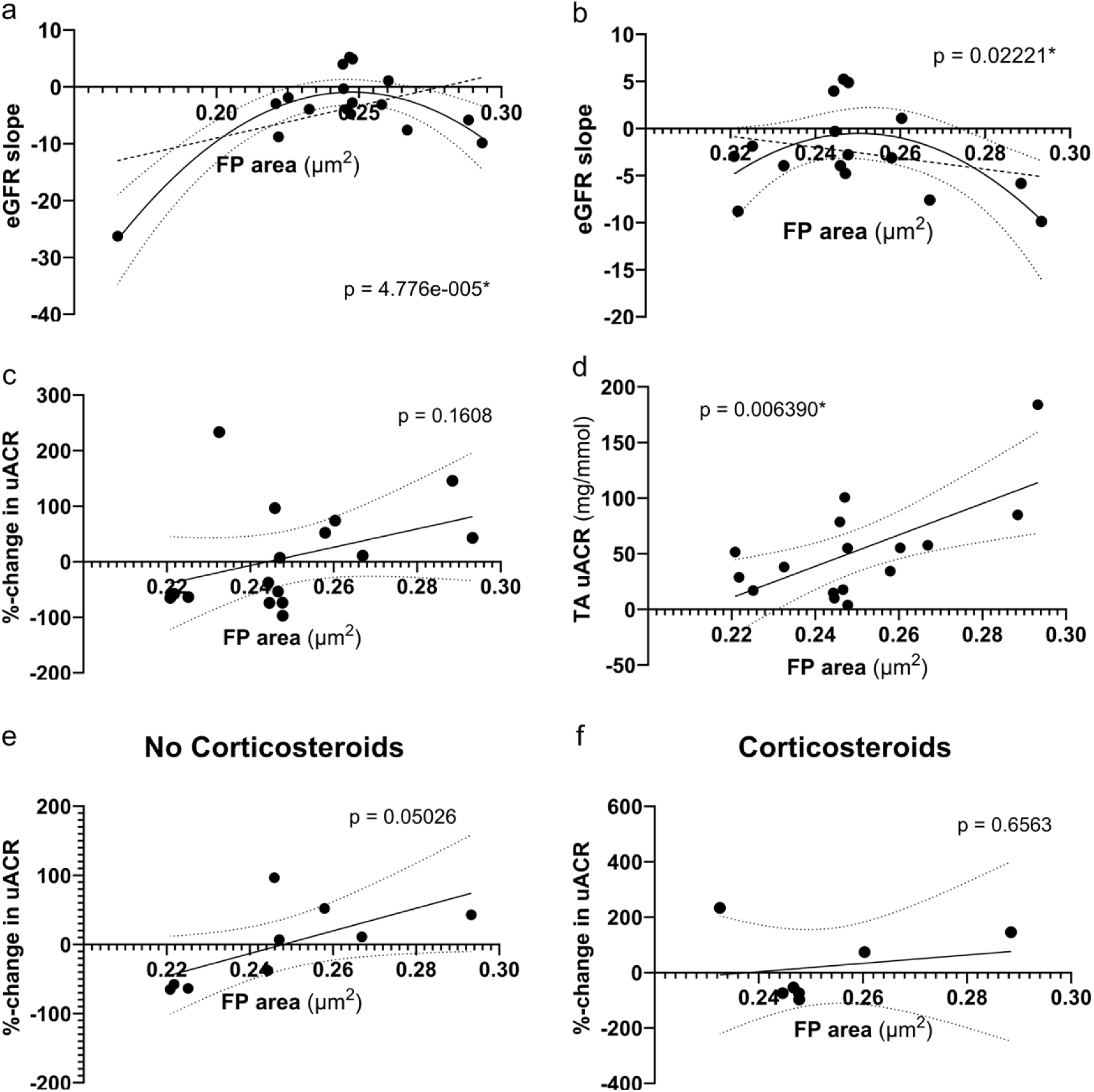
FP area correlates with eGFR slope and long-term albuminuria in IgA Nephropathy. The graphs show scatterplots with regression lines for the relationship between FP area and eGFR slope, %-change in uACR and TA uACR. Each point represents the mean value for one patient. eGFR slope was measured as the yearly change in eGFR (mL/min/1.73m^2^/year). (a, b) The correlation between FP area and eGFR slope was tested by fitting both a quadratic polynomial regression and a linear regression, both of which are shown in the graphs. The fit of the polynomial regression was compared with the linear regression using an F-test. For the other graphs, only simple linear regression was used for statistical analysis (c-f). (a) FP area had an inverse U-shape relationship with eGFR slope where both low and high FP area was associated with worse eGFR slope. (b) The relationship remained significant after removal of the most negative outlier indicating robust correlation. (c-f) The patient with the lowest FP area, seen in Figure 5a, was not included in the correlative analyses with %-change in uACR and TA uACR due to having less than three uACR measurements. (c, d) FP area was not correlated with %-change in uACR (c) but higher FP area was significantly correlated with higher TA uACR (d). In corticosteroid subgroups, there was a non-significant trend for higher FP area to be correlated with increasing uACR during the first year among untreated patients (e) which was not seen with corticosteroid treatment (f).

#### High FP circularity is associated with improvement in uACR during the first year in IgAN

FP circularity was not correlated with eGFR slope in either the total IgAN cohort (Figure 6a) or within the subgroups stratified by CS treatment status (data not shown). Moreover, FP circularity did not correlate to baseline uACR but higher FP circularity was associated with improvement in uACR during the 1st year (Figure 6b). This trend toward improved uACR was consistent in both CS subgroups but did not reach statistical significance (Figure 6c-d). There was also a non-significant trend suggesting an association between higher FP circularity and lower TA uACR (Figure 6e).

**Figure 6.**
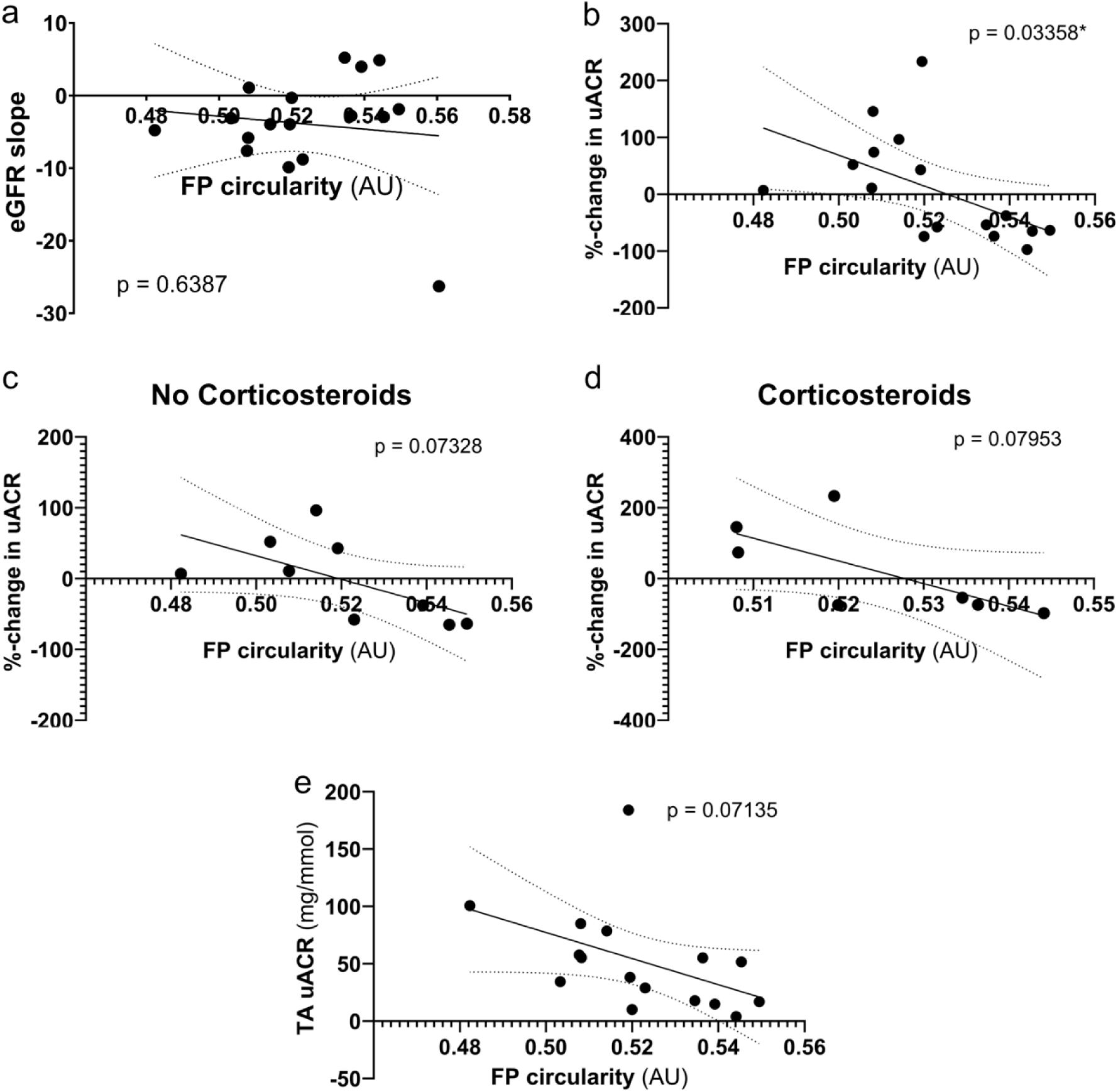
High FP circularity correlates with improvement in uACR during the first year in IgA Nephropathy. The graphs show scatterplots with fitted linear regressions for the relationship between FP area and eGFR slope (a), %-change in uACR (c, d, e) and TA uACR (d). Subgroup analysis stratified by if the patient received corticosteroids during follow-up was also performed for %-change in uACR (c, d). Each point represents the mean value for one patient. eGFR slope was measured as the yearly change in eGFR (mL/min/1.73m^2^/year). Correlations were tested by linear regression analysis. (a) FP circularity did not correlate with eGFR slope in IgAN. (b) Higher FP circularity was associated with a reduction in uACR in IgAN during the first year. All patients with a mean FP circularity exceed 0.53 saw significant improvement in uACR. (c, d) In corticosteroid subgroups, a non-significant trend toward higher FP circularity being associated with reduction in uACR remained among both untreated and treated IgAN patients. (e) There was a non-significant trend towards higher FP circularity being associated with lower TA uACR mirroring the improvement in uACR among patients with high FP circularity. *AU = Arbitrary units*.

#### Combined morphometric parameters as predictors of clinical outcome in IgAN

Given that the different morphometric features correlated differently to clinical parameters, we wanted to explore if any combination of morphometric features could better identify progressive disease in IgAN. For visual clarity due to the large spread in eGFR slope, patients were stratified into eGFR slope quartiles with the first quartile having the most negative slope and the fourth quartile having the most positive slope.

Bivariate analysis of SDL versus FP circularity revealed a tendency for the most negative quartiles of eGFR slope separating towards lower SDL and FP circularity (Suppl. Figure 4a). To corroborate this, we calculated the SDL-FP circularity product which proved to be a more significant predictor of eGFR slope than SDL alone (p = 0.021 vs p = 0.041).

Examination of the other combinations of SDL, FP area & FP circularity confirmed that the steepest eGFR declines occurred at the extremes of small and large FP area, while higher FP circularity indicated a better prognosis (Suppl. Figure 4c-d).

### Incidental finding of podocytes on Bowman’s capsule

As an incidental finding of applying high-resolution 3D microscopy, we found 2 patients (one with IgAN and one with LN) that had podocytes on Bowman’s capsule with organized slit diaphragm networks (Supplemental Figure 5). These two cases were found among the 11 cases that had been co-stained with collagen IV where Bowman’s capsule could be differentiated.

## Discussion

The principal finding of this study is that nanoscale podocyte morphometrics, quantified by deep learning-based segmentation of high-resolution confocal images, identify associations with disease progression and treatment response in IgAN that are not captured by conventional electron microscopy. These results suggest that quantitative FP analysis may provide clinically actionable information beyond current biopsy evaluation. Importantly, the morphometric parameters were measured by automated analysis and therefore independent of individual assessment bias.

SDL, a validated indicator of FP effacement,^13,21^ correlated significantly with uACR in our cohort, whereas conventional EM-based assessment did not. This difference likely reflects the inherent advantage of analyzing the entire capillary surface in three dimensions rather than relying on cross-sectional EM images, which sample only a fraction of the glomerular filtration area. Interestingly, lower SDL was associated with a steeper eGFR decline in IgAN patients. In subgroup analysis stratified by corticosteroid therapy, this association persisted as a trend in untreated patients (p = 0.068) but was absent in the treated group, raising the possibility that corticosteroids mitigate the risk of progression associated with severe FP effacement. This interpretation is consistent with recent evidence that podocyte lesions in IgAN respond favorably to corticosteroid therapy and that FSGS-type lesions predict treatment response.^15^ Mechanistically, we have previously shown in mouse models that low SDL leads to podocyte loss,¹³ which could represent the pathophysiological link between severe effacement and declining kidney function. A key challenge in implementing corticosteroid therapy guided by podocyte injury is the current lack of objective, numerical and reproducible measurements. SDL could address this gap as a quantitative marker, but our findings require validation in adequately powered cohorts before such an application can be considered.

FP area instead showed an inverse U-shape relationship with eGFR slope in IgAN where both low and high FP area were associated with a more negative slope, potentially due to different mechanisms. Higher FP area was also strongly correlated with higher TA uACR, higher eGFR and lower GSG%. We hypothesize that an increase in FP area represents an early sign of podocyte injury that precedes overt clinical deterioration, and that patients with this morphometric profile may be undertreated under current management strategies. If confirmed, this finding could have direct implications for treatment decisions in patients with apparently mild IgAN.

FP circularity was associated with short-term changes in albuminuria. Higher FP circularity at biopsy predicted improvement in uACR during the first year. Of 16 IgAN patients with available follow-up, six of eight whose uACR improved had FP circularity above 0.53. None of the eight patients with worsening uACR had FP circularity above this threshold. When combined with SDL or FP area, higher circularity also tended to predict a more favorable eGFR trajectory. In mouse models, we have previously shown that circularity increases early in both genetic and inflammatory injury, while SDL decreases later in the disease course.¹³ FP circularity may therefore reflect an early, reversible stage of podocyte injury, possibly related to hyperfiltration, that improves even with conservative management.

These observations argue against relying on any single morphometric parameter to characterize podocyte disease. Assessment of podocyte alterations has historically focused on FP effacement, largely due to the limitations of cross-sectional EM imaging. Our data indicate that the morphological response of podocytes to injury is more complex than effacement alone. In our previous study in MCD, SDL and FP circularity varied independently, and FP circularity was higher among pediatric patients, suggesting that it reflects an earlier or mechanistically distinct phase of injury.²¹ In the same study, two mouse models with comparable SDL could only be distinguished by their FP circularity.²¹ Future studies should therefore incorporate multiple morphometric parameters. With further mechanistic work, it may become possible to link specific morphometric profiles to underlying disease pathways and inform individualized treatment decisions. The deep learning tool AMAP had previously been trained and validated on FFPE sections across several glomerular diseases including MCD, FSGS and IgAN.²¹ Here, we used frozen sections to preserve tissue for biomarker studies currently in progress. The original model required image preprocessing and fine-tuning to accommodate the lower signal-to-background ratio of frozen-section preparation. Both visual and morphometric results were nonetheless consistent with our previous FFPE-based analyses, indicating that the approach can be applied across different tissue preparation methods.

The principal limitation of this study is the small cohort (n = 37 overall, n = 19 for IgAN), which limits statistical power and precludes multivariable adjustment. Subgroup analyses stratified by treatment are therefore exploratory. The number of glomeruli available for morphometric analysis also varied between patients, introducing sampling variability. That morphometric parameters showed significant or near-significant associations with surrogate endpoints of kidney disease progression, eGFR slope, time-averaged uACR and change in uACR, despite these constraints speaks to the sensitivity of the method. Validation in larger, multicenter cohorts with standardized tissue preparation and prospective follow-up is needed to confirm these associations and establish clinically useful thresholds.

## Conclusion

Nanoscale podocyte morphometrics provide a more sensitive and comprehensive assessment of podocyte injury than conventional biopsy evaluation. In this proof-of-concept study in IgAN, distinct morphometric parameters were associated with kidney disease progression, albuminuria trajectory and potential treatment response. If validated, this approach could enable objective, reproducible quantification of podocyte injury for risk stratification and treatment guidance, addressing an unmet need in the clinical management of glomerular disease.

## Data Availability

Due to privacy laws and ethical restrictions concerning pseudonymized patient data under the EU General Data Protection Regulation (GDPR) and Swedish law, the clinical data and images underlying this study cannot be made publicly available. Data may be shared with qualified researchers upon reasonable request to the corresponding author after establishment of a formal data transfer agreement and approval from the Swedish Ethical Review Authority.

## Acknowledgments

We acknowledge microscopy support from the Advanced Light Microscopy (ALM) Facility, Royal Institute of Technology (KTH), SciLifeLab Solna, Sweden and the National Microscopy Infrastructure, NMI (VR-RFI 2023-00163). The work was supported by grants to HB from the Clinical Technology Development Project at SciLifeLab. RE and DUJ were funded by the Torsten Söderbergs Stiftelse, Stockholm, Sweden. RE were also funded by “Stiftelsen Stig och Gunborg Westman” for research on kidney disease, transplantation and organ donation. RE was supported through funding and clinical fellowship with AIDA, Analytic Imaging Diagnostics Areana, Linköping, Sweden. DUJ was supported by funding from University of Cologne, Germany. TB is supported by the German Research Foundation, TRR/CRU 422, project A1.

## Statement of Ethics

Ethical approval for the BIONEF DS project has been received by the Swedish Ethical Review Authority with Dnr 2018/803-31 with approved amendments Dnr 2023-01265-02, Dnr 2024-01876-02, and and Dnr 2025-05680-02 pertaining to the present study. Written informed consent was obtained from all subjects at recruitment. The study was conducted in accordance with the Declaration of Helsinki.

## References

1. Kidney Disease: Improving Global Outcomes (KDIGO) Glomerular Diseases Work Group. KDIGO 2021 Clinical Practice Guideline for the Management of Glomerular Diseases. Kidney Int. 2021;100(4S):S1–S276. doi:10.1016/j.kint.2021.05.021

2. Working Group of the International IgA Nephropathy Network and the Renal Pathology Society, Cattran DC, Coppo R, et al. The Oxford classification of IgA nephropathy: rationale, clinicopathological correlations, and classification. Kidney Int. 2009;76(5):534–545. doi:10.1038/ki.2009.243

3. Trimarchi H, Barratt J, Cattran DC, et al. Oxford Classification of IgA nephropathy 2016: an update from the IgA Nephropathy Classification Working Group. Kidney Int. 2017;91(5):1014–1021. doi:10.1016/j.kint.2017.02.003

4. Bajema IM, Wilhelmus S, Alpers CE, et al. Revision of the International Society of Nephrology/Renal Pathology Society classification for lupus nephritis: clarification of definitions, and modified National Institutes of Health activity and chronicity indices. Kidney Int. 2018;93(4):789–796. doi:10.1016/j.kint.2017.11.023

5. Barbour SJ, Espino-Hernandez G, Reich HN, et al. The MEST score provides earlier risk prediction in lgA nephropathy. Kidney Int. 2016;89(1):167–175. doi:10.1038/ki.2015.322

6. The IgA Nephropathy Study Group in Japan, Hisano S, Joh K, et al. Reproducibility for pathological prognostic parameters of the Oxford classification of IgA nephropathy: a Japanese cohort study of the Ministry of Health, Labor and Welfare. Clin Exp Nephrol. 2017;21(1):92–96. doi:10.1007/s10157-016-1258-8

7. Bellur SS, Roberts ISD, Troyanov S, et al. Reproducibility of the Oxford classification of immunoglobulin A nephropathy, impact of biopsy scoring on treatment allocation and clinical relevance of disagreements: evidence from the VALidation of IGA study cohort. Nephrol Dial Transplant. 2019;34(10):1681–1690. doi:10.1093/ndt/gfy337

8. Testa F, Fontana F, Pollastri F, et al. Automated Prediction of Kidney Failure in IgA Nephropathy with Deep Learning from Biopsy Images. Clin J Am Soc Nephrol. 2022;17(9):1316–1324. doi:10.2215/CJN.01760222

9. Jaugey A, Maréchal E, Tarris G, et al. Deep learning automation of MEST-C classification in IgA nephropathy. Nephrol Dial Transplant. 2023;38(7):1741–1751. doi:10.1093/ndt/gfad039

10. Hölscher DL, Bouteldja N, Joodaki M, et al. Next-Generation Morphometry for pathomics-data mining in histopathology. Nat Commun. 2023;14(1):470. doi:10.1038/s41467-023-36173-0

11. Huo Y, Deng R, Liu Q, Fogo AB, Yang H. AI applications in renal pathology. Kidney Int. 2021;99(6):1309–1320. doi:10.1016/j.kint.2021.01.015

12. Benzing T, Salant D. Insights into Glomerular Filtration and Albuminuria. N Engl J Med. 2021;384(15):1437–1446. doi:10.1056/NEJMra1808786

13. Butt L, Unnersjö-Jess D, Höhne M, et al. A molecular mechanism explaining albuminuria in kidney disease. Nat Metab. 2020;2(5):461–474. doi:10.1038/s42255-020-0204-y

14. Bellur SS, Lepeytre F, Vorobyeva O, et al. Evidence from the Oxford Classification cohort supports the clinical value of subclassification of focal segmental glomerulosclerosis in IgA nephropathy. Kidney Int. 2017;91(1):235–243. doi:10.1016/j.kint.2016.09.029

15. Bellur SS, Troyanov S, Vorobyeva O, Coppo R, Roberts ISD, Validation in IgA Nephropathy study group. Evidence from the large VALIGA cohort validates the subclassification of focal segmental glomerulosclerosis in IgA nephropathy. Kidney Int. 2024;105(6):1279–1290. doi:10.1016/j.kint.2024.03.011

16. Unnersjö-Jess D, Scott L, Blom H, Brismar H. Super-resolution stimulated emission depletion imaging of slit diaphragm proteins in optically cleared kidney tissue. Kidney International. 2016;89(1):243–247. doi:10.1038/ki.2015.308

17. Unnersjö-Jess D, Butt L, Höhne M, et al. A fast and simple clearing and swelling protocol for 3D in-situ imaging of the kidney across scales. Kidney Int. 2021;99(4):1010–1020. doi:10.1016/j.kint.2020.10.039

18. Siegerist F, Ribback S, Dombrowski F, et al. Structured illumination microscopy and automatized image processing as a rapid diagnostic tool for podocyte effacement. Sci Rep. 2017;7(1):11473. doi:10.1038/s41598-017-11553-x

19. Pullman JM. New Views of the Glomerulus: Advanced Microscopy for Advanced Diagnosis. Front Med (Lausanne). 2019;6:37. doi:10.3389/fmed.2019.00037

20. Ali A, Liu Z, Ye K, et al. Super-resolved optical imaging, reconstruction, and spatial analysis of whole mouse glomeruli via the Glomerulus Mapping and Analysis Pipeline. Kidney Int. 2025;108(5):901–910. doi:10.1016/j.kint.2025.07.024

21. Unnersjö-Jess D, Butt L, Höhne M, et al. Deep learning-based segmentation and quantification of podocyte foot process morphology suggests differential patterns of foot process effacement across kidney pathologies. Kidney Int. 2023;103(6):1120–1130. doi:10.1016/j.kint.2023.03.013

22. Siegerist F, Hay E, Hammer E, et al. The differential expression of MAGI2 in glomerulopathies and its application as a molecular discriminator of podocytopathies. J Transl Med. 2025;23:701. doi:10.1186/s12967-025-06696-9

23. Schindelin J, Arganda-Carreras I, Frise E, et al. Fiji: an open-source platform for biological-image analysis. Nat Methods. 2012;9(7):676–682. doi:10.1038/nmeth.2019

24. Ronneberger O, Fischer P, Brox T. U-Net: Convolutional Networks for Biomedical Image Segmentation. arXiv. Preprint posted online May 18, 2015:arXiv:1505.04597. doi:10.48550/arXiv.1505.04597

25. Haaskjold YL, Lura NG, Bjørneklett R, Bostad LS, Knoop T, Bostad L. Long-term follow-up of IgA nephropathy: clinicopathological features and predictors of outcomes. Clin Kidney J. 2023;16(12):2514–2522. doi:10.1093/ckj/sfad154

