## Supplementary Information: Methods, Figures & Tables for "Nanoscale Podocyte Morphometrics Predict Disease Progression in IgA Nephropathy"

#### Supplementary Methods:

##### Patient cohort:

The 37 patients in this study were selected from the BIONEF-DS study cohort, a patient cohort from the Nephrology Clinic at Danderyd University Hospital, Stockholm, Sweden, that has been built up since 2019 for the study of biomarkers in glomerular disease. For this study, all patients undergoing diagnostic kidney biopsy were asked for participation, including their agreement to collect remaining biopsy material, blood and urine samples as well as prospective clinical data. Kidney biopsies had been stored in the Stockholm medical biobank at the Department of Pathology, Karolinska University Hospital, Stockholm.

For the present study, participants were selected from the cohort based on having an available biopsy and being diagnosed with the immune-mediated glomerular diseases IgA Nephropathy, IgA Vasculitis, Membranous nephropathy or Lupus nephritis. As comparators, two cases of hereditary basement membrane defect were selected to represent non-immune-mediated glomerular disease and three cases of nephrosclerosis as regular controls. Patients were selected sequentially based on follow-up time with the only exception being prioritizing selecting patients with IgA Nephropathy to make up at least half the cohort for the present study.

##### Sample preparation:

The biopsies analyzed in this study were stored fresh-frozen in OCT. After thawing, the biopsy was fixated in PFA 4% for either 1h in RT or overnight in 4°C. The biopsy was sectioned by vibratome and cleared for either 1h or 24h at 50°C depending on fixation time using 4% SDS w/ Boric acid according to the protocol by David Unnersjö-Jess et al.[17]

Immunolabelling was performed using a primary anti-nephrin antibody (R&D Systems, cat. no. AF4269) diluted to 1:100 and a secondary antibody with an Alexa Fluor 488 dye (Thermo Fisher Scientifics cat. no. A20000) or an Alexa Fluor 405 dye (Abcam cat no. ab175649) diluted to 1:100. Dilutions were made in HEPES-TCS buffer w/ 8% Triton X-100. 11 of the biopsies were co-stained for Collagen IV using a primary antibody (Abcam EPR22911-127) conjugated to Alexa Fluor 555 in-house (Thermo Fisher Scientifics cat. no. A37571). After initial image analysis on the first batch of images, collagen IV was determined to not be necessary for the analysis of podocyte morphology as AMAPs nephrin-based ROI selection was robust. Therefore, the remaining biopsies were not co-stained for collagen IV.

Before imaging, the sample was embedded in a saturated fructose-solution (80.2% w/w in DI-H2O) and mounted on a MatTek (MatTek, Ashland, MA) or Ibidi (Ibidi GmBH, Gräfelfing, Germany) imaging dish. Imaging was carried out on a Leica TCS SP8 confocal microscope using a 100X/1.4 NA oil immersion objective (Leica Microsystems, Buffalo Grove, IL) with the pinhole set to 0.3 Airy units.

#### Podocyte morphometric parameters:

Slit diaphragm length (SDL) was calculated by measuring the length of the slit diaphragm and then dividing by the capillary surface area covered by the slit diaphragm (SD) for a standardized measurement. The area to be divided with was automatically assigned based on the SD stain as described in the original AMAP publication.[21] FP area was calculated as the area of each FP instance segmentation and FP perimeter as the circumference of the same segmentation. FP circularity was calculated using the formula; circularity = 4 π (area/perimeter^2).

#### Image pre-processing:

Roughly 20% of the 439 raw images required additional pre-processing for successful segmentation by the model. The 2 main image aberrations affecting segmentation quality were low absolute intensities and noise. To mitigate issues with low intensities, 69 images (15.7%) were improved through normalization using Fiji:s auto adjust brightness/contrast function and 11 images (2.5%) using local contrast enhancement (CLAHE in Fiji). For issues with noise, 21 images (4.8%) were improved using deconvolution in Huygens software utilizing Classic Maximum Likelihood Estimation for 15 images (3.4%) and Classical Tikhonov-Miller deconvolution for 6 images (1.4%) (Scientific Volume Imaging, The Netherlands, http://svi.nl). All images with pre-processing were included in the final analysis.

#### Fine-tuning of the model:

The original pre-trained model checkpoint was subjected to a two-step fine-tuning process. Initially, the model was fine-tuned on four annotated samples for 100 epochs with a learning rate of 0.001. Subsequently, it was further fine-tuned on nine additional samples for 200 epochs, employing a learning rate of 0.001. Given the limited availability of annotated samples, the model's performance was evaluated on the original AMAP test dataset throughout the fine-tuning stages. Furthermore, the qualitative performance of the fine-tuned checkpoints on unseen sample types was assessed by expert evaluation.

For image analysis, the AMAP model, based on an altered U-Net architecture was initially applied to all images using its original publication weights. This initial application revealed limitations, particularly in segmenting slit and foot processes in several images (e.g., Figure OLD_AMAP_A). To address these challenges, the model underwent a two-step fine-tuning process. The first fine-tuning step involved training on four annotated images from the new image types, which led to a notable improvement in overall segmentation. However, the model continued to exhibit difficulties with more severely diseased samples. Consequently, a second fine-tuning step was performed, starting from the previously fine-tuned checkpoint, utilizing nine additional annotated images representing advanced disease states. During both fine-tuning phases, model performance was continuously monitored against the original AMAP test dataset, and qualitative assessments by experts were conducted on unseen sample types.

### Supplemental Figures

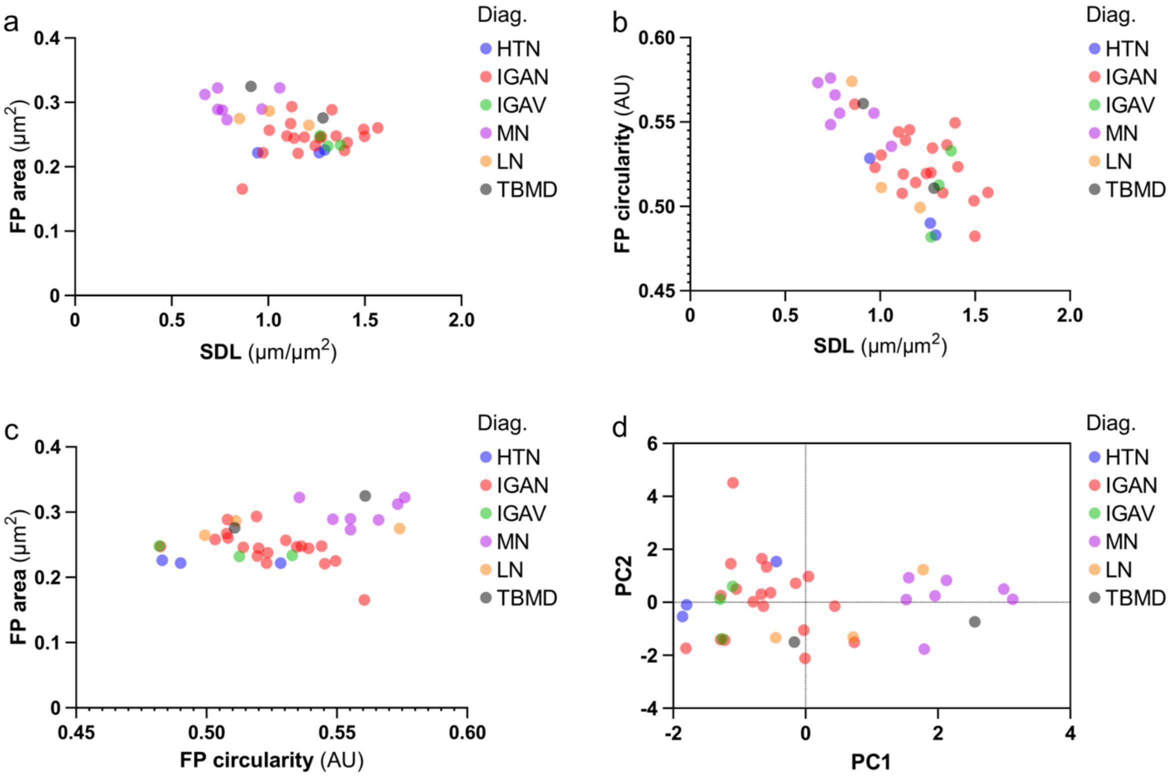


Supplemental Figure 1. - **Cross-correlations and principal component analysis of morphometric features across diagnoses.** The graphs show scatterplots of the podocyte morphometric features in correlation to each other and principal component analysis. Each dot represents the mean value for one patient. (a, b, c) Scatterplots showing the relationship between the morphometric features SDL, FP area and FP circularity in the whole cohort. Broadly, there was a tendency for cross-correlations between all morphometric features across diseases but with significant variation between and within diagnosis groups. (d) Scatterplot of PC1 versus PC2 selected by principal component analysis using the morphometric parameters SDL, FP circularity, FP perimeter and FP area. The graph shows separation into 2 broad clusters. The right cluster includes all MN-patients, one patient with LN (classified as Membranous LN, WHO 5) and one with TBMD. The rest of the patients associated towards the left.

*AU = Arbitrary units.*

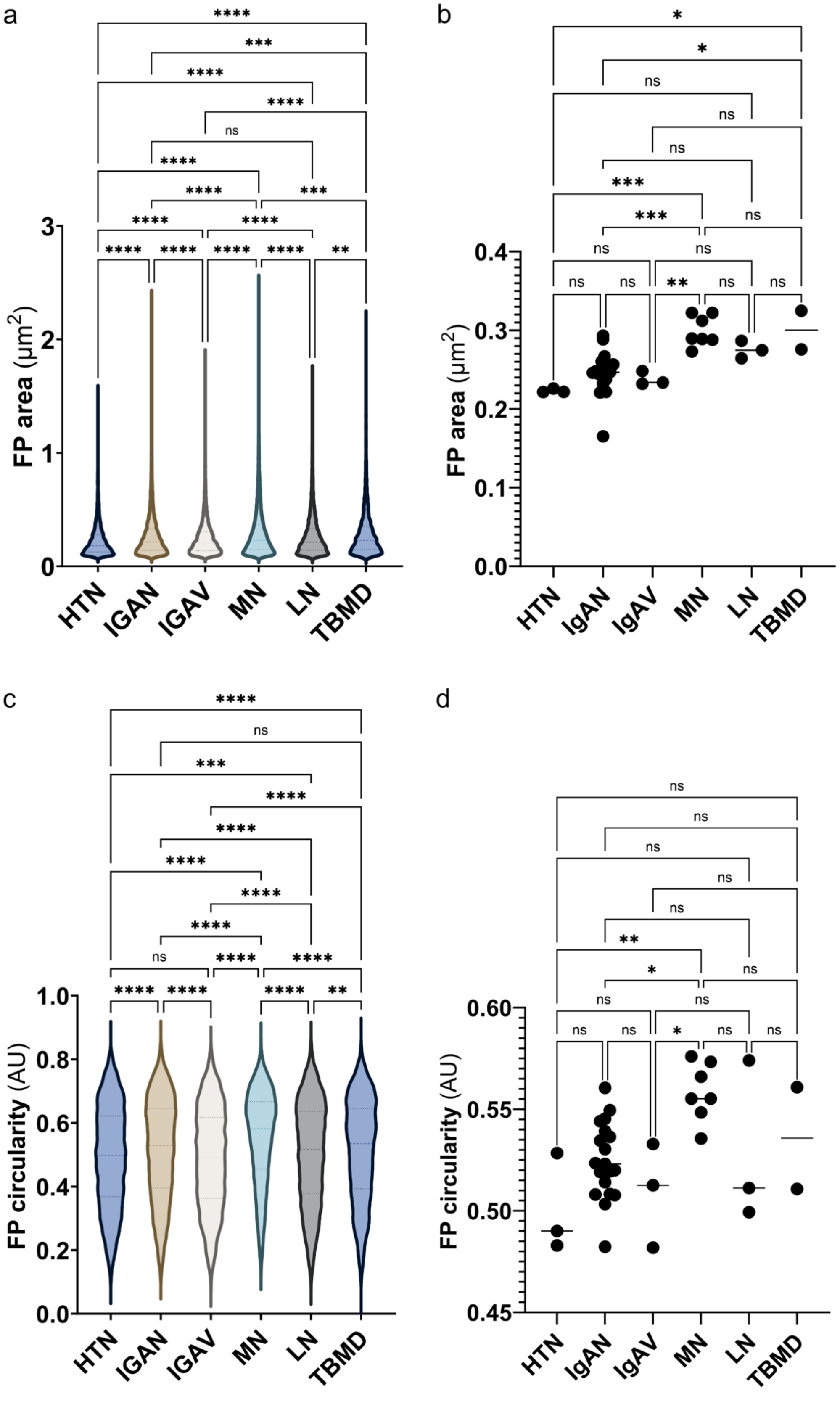
a
FP area (μm2)
3
2
1
0
HTN
IGAN
IGAV
MN
LN
TBMD
b
FP area (μm2)
0.4
0.3
0.2
0.1
0.0
HTN
IgAN
IgAV
MN
LN
TBMD
c
FP circularity (AU)
1.0
0.8
0.6
0.4
0.2
0.0
HTN
IGAN
IGAV
MN
LN
TBMD
d
FP circularity (AU)
0.60
0.55
0.50
0.45
HTN
IgAN
IgAV
MN
LN
TBMD

Supplemental Figure 2. - **Comparison of distribution and mean foot process morphometrics between diagnoses.** The violin plots show the distribution of FP area (a) and FP circularity (c) for all individual segmented FPs, grouped by diagnosis. The strip plots show comparisons of the mean FP area (b) and FP circularity (d) between diagnoses where each dot represents the mean of one patient. (a, b) FP area showed significantly different distribution between most diagnoses (a) with the most significant differences in mean between MN and HTN, IgAN and IgAV (b). (c, d) FP circularity showed a similar pattern of differing distribution between most diagnoses (c) and different mean between MN and HTN, IgAN and IgAV (d). Despite the differences in distribution between diagnoses, major similarities, especially in the distribution peak, is still seen for both FP area (a) and FP circularity (c).
*AU = Arbitrary units.*

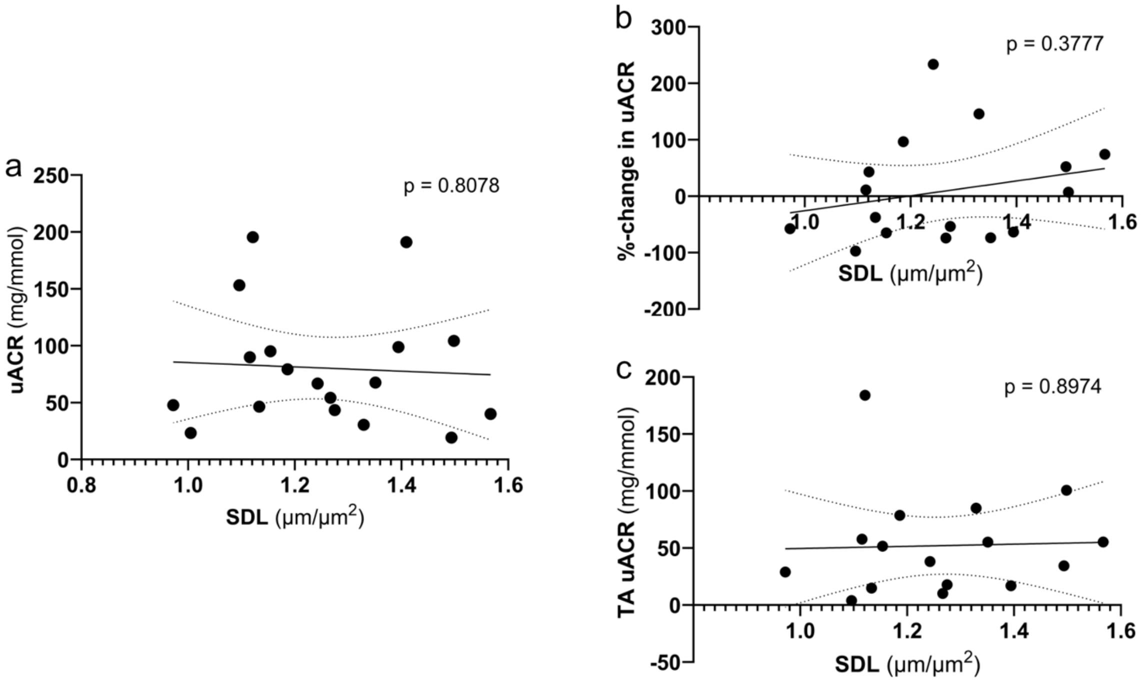


Supplemental Figure 3 - **Correlation of SDL with uACR, %-change in uACR during the first year and TA uACR.** The graphs show scatterplots with a fitted linear regression for the relationship between SDL and uACR (a), %-change in uACR (b) and TA uACR (c). Each point represents the mean value of SDL for one patient. (a, b, c) Correlations were tested by linear regression analysis. There was no correlation between SDL and uACR (a), %-change in uACR (b) or TA uACR (c)

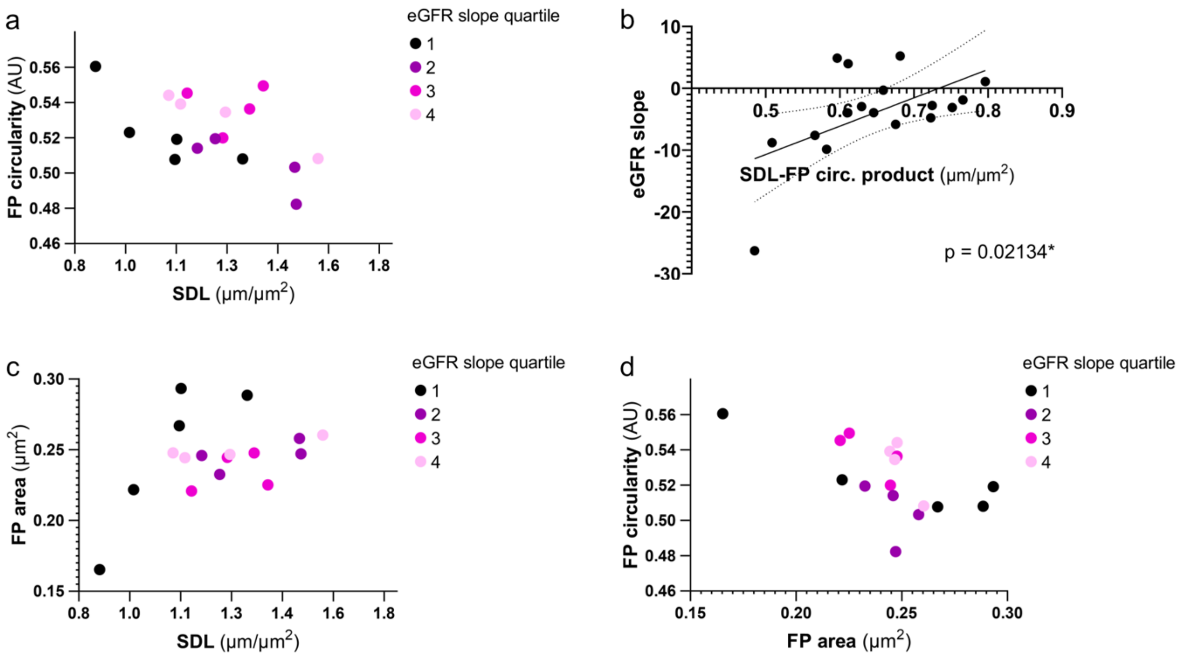


Supplemental Figure 4 - **Correlations between combinations of podocyte morphometric parameters and eGFR slope grouped by quartiles.** The figure shows scatterplots of morphometric parameters where each point represents the mean value for one patient. Each point is colored by the quartile of eGFR slope where the first quartile has the most negative slope (black) and the fourth quartile has the most positive slope (light pink). eGFR slope was measured as the yearly change in eGFR (mL/min/1.73m$^2$/year). (a) Plotting SDL versus FP circularity there was tendency for a diagonal separation where the most negative quartiles separate toward the bottom-left with low SDL and low FP circularity relative to degree of SDL. (b) To confirm this finding, the SDL-FP circularity product was calculated for each patient and tested for correlation with the eGFR slope by linear regression analysis. The SDL-FP circularity product correlated significantly with the eGFR slope and was a more significant predictor than SDL alone ($p = 0.021$ vs $p = 0.041$) (Results for SDL alone is available in Fig 4a.) (c) For SDL versus FP area, the patients in the most negative quartile all have either very low SDL or very high FP area. (d) Plotting FP area versus FP circularity, the patients in the most negative eGFR quartile had either very low or very high FP area. Among intermediate values for FP area, a higher FP circularity tended to be associated with a better eGFR slope.
*AU = Arbitrary units.*

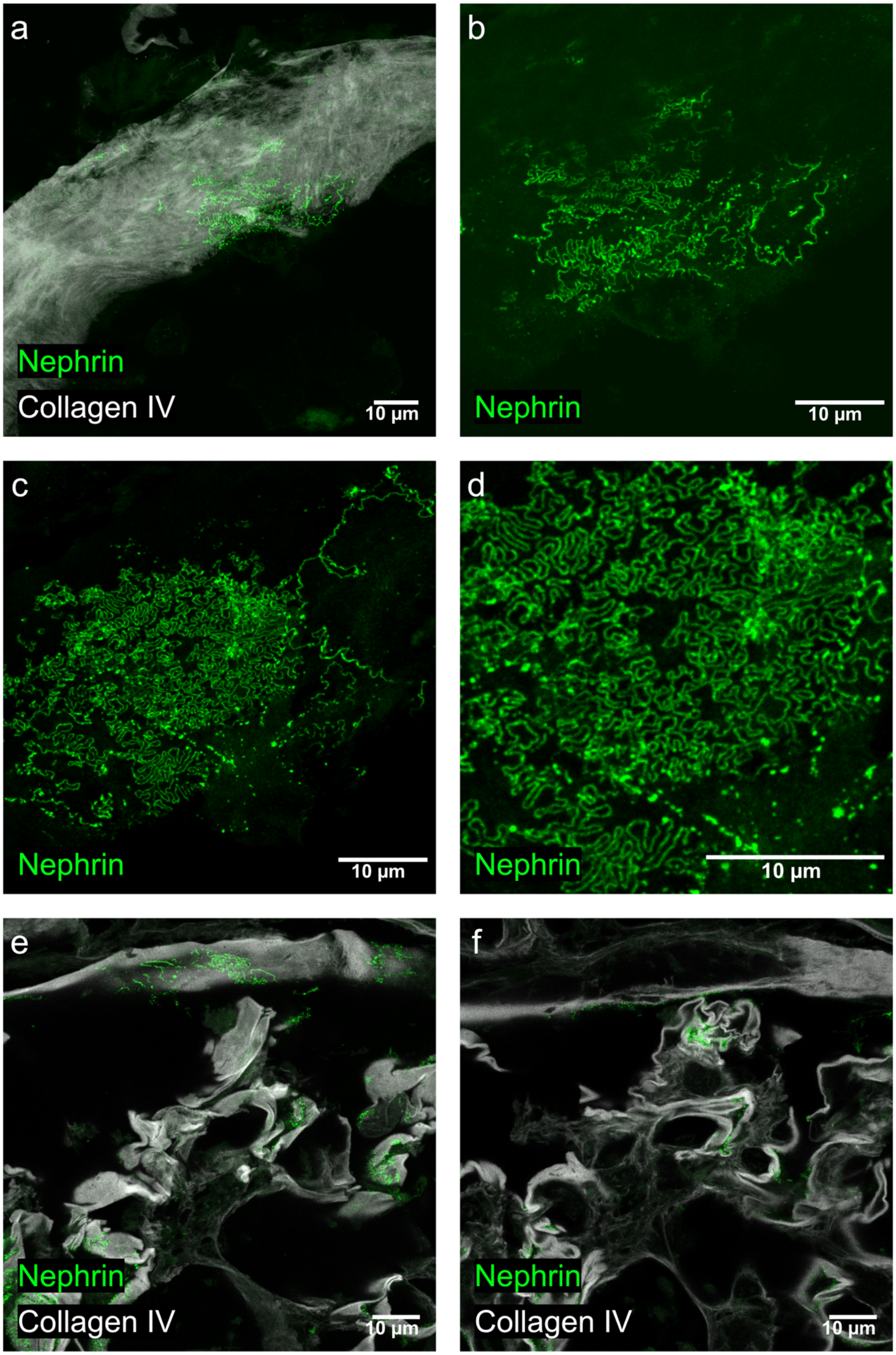

a
Nephrin
Collagen IV
10 µm
b
Nephrin
10 µm
c
Nephrin
10 µm
d
Nephrin
10 µm
e
Nephrin
Collagen IV
10 µm
f
Nephrin
Collagen IV
10 µm

Supplement Figure 5 – **Incidental finding of podocytes with intact foot process architecture on Bowman's capsule in SLE and IgAN.** (a, b, c, d) Confocal images of a podocyte on Bowman's capsule in a patient with SLE, dual-stained for nephrin and collagen IV. The image stacks have been maximum intensity projected. (a) The image shows the podocyte being attached to the Bowman's capsule with the typical weave-like collagen IV structure. (b) Zoomed in image of the same podocyte with the collagen IV-channel removed shows how the nephrin network branches out from the cell body of the podocyte. (c) Deeper images of the same podocyte show how the nephrin forms a large nest-like structure in the center with a less organized structure towards the right-hand side. (d) Zooming in at the same deeper sections reveals formed foot process-like structures in the slit diaphragm network on the Bowman's capsule. (e) The image shows slit formation on Bowman's capsule in a different patient with IgAN. In the top of the image the slit diaphragm network can be seen covering the Bowman's capsule in an en-face view. Next to the slit diaphragm network, a collapsing capillary tuft is seen. (f) Imaging deeper in the same area reveals that the nephrin signal stretches further along Bowman's capsule, approximately 30-50 micrometers left-ward away from the nearby tuft.

### Supplemental Tables

| | No Immunosuppression | Immunosuppression | P-Value |
|---|---|---|---|
| **Clinical parameters** | | | |
| **Age, years (median [IQR])** | 41.5 [35.5-45.5] | 52.0 [34.0-58.0] | 0.525† |
| **Sex (Male:Female, %)** | 8:4, 66.7% | 7:0, 100% | 0.245 |
| **eGFR, mL/min/1.73m2 (mean ± SD)** | 65.7 ± 29.5 | 62.1 ± 23.2 | 0.771 |
| **MAP, mmHg (mean ± SD)** | 98.0 ± 8.2 | 96.4 ± 8.4 | 0.690 |
| **BMI, kg/m2 (median [IQR])** | 24.0 [23.3-27.2] | 24.9 [24.1-29.8] | 0.299† |
| **uACR, mg/mmol (median [IQR])** | 92.4 [47.4-125.9] | 54.2 [41.7-67.2] | 0.227† |
| **Hypertension** | 9/12 (75.0%) | 5/7 (71.4%) | 1.000 |
| **Histological parameters** | | | |
| **GSG% (median [IQR])** | 27.2 [19.2-62.1] | 7.5 [5.8-28.8] | 0.108† |
| **Oxford M** | 10/12 (83.3%) | 7/7 (100.0%) | 0.509 |
| **Oxford E** | 6/12 (50.0%) | 5/7 (71.4%) | 0.633 |
| **Oxford S** | 11/12 (91.7%) | 5/7 (71.4%) | 0.523 |
| **Oxford T** | 0.5 [0.0-1.2] | 0.0 [0.0-1.0] | 0.710† |
| **Oxford C** | 0.0 [0.0-0.0] | 1.0 [0.0-1.0] | 0.027*† |
| **Morphometric features** | | | |
| **SDL, μm/μm$^2$ (mean ± SD)** | 1.2 ± 0.2 | 1.3 ± 0.1 | 0.198 |
| **FP area, μm$^2$ (mean ± SD)** | 0.2 ± 0.0 | 0.3 ± 0.0 | 0.288 |
| **FP circularity, AU (mean ± SD)** | 0.5 ± 0.0 | 0.5 ± 0.0 | 0.959 |

Supplemental Table 1 - **Correlation between baseline parameters and morphometric features with the decision to treat with immunosuppression during follow-up in IgAN.** The table shows the correlations between baseline clinical parameters, histological parameters and morphometric features with the decision to treat with immunosuppression during follow-up in IgAN. The continuous variables were tested for normality by Shapiro-Wilk test. Mean ± SD is presented for normally distributed variables and median with interquartile range (IQR) is present for non-normally distributed variables. For normally distributed variables, parametric Student's T-test was performed. For non-normal variables, non-parametric Mann-Whitney U-test was performed. All results from the non-parametric test are marked with "†". For categorical variables, Fisher's exact test was used. Positive p-values are denoted with "*". *AU = Arbitrary units.*

| Diagnosis | Mean mode | CV% | |
|---|---|---|---|
| **Total Cohort** | 0.153 | 7.93 | |
| **HTN** | 0.133 | 2.72 | |
| **IGAN** | 0.152 | 5.55 | |
| **IGAV** | 0.149 | 9.23 | |
| **MN** | 0.159 | 7.30 | |
| **LN** | 0.158 | 12.37 | |
| **TBMD** | 0.161 | 11.29 | |
| **Comparison of the mean mode for FP area between diagnosis groups** | | | |
| **Group** | **Comparison Group** | **Mean difference** | **Adj. P-value** |
| **HTN** | IGAN | 0.0189 | 0.0788 |
| **HTN** | IGAV | 0.0159 | 0.4734 |
| **HTN** | MN | 0.0258 | 0.0176* |
| **HTN** | LN | 0.0248 | 0.0809 |
| **HTN** | TBMD | 0.0275 | 0.0845 |
| **IGAN** | IGAV | -0.003 | 0.9975 |
| **IGAN** | MN | 0.0069 | 0.7018 |
| **IGAN** | LN | 0.0058 | 0.95 |
| **IGAN** | TBMD | 0.0086 | 0.8892 |
| **IGAV** | MN | 0.0099 | 0.7671 |
| **IGAV** | LN | 0.0089 | 0.9121 |
| **IGAV** | TBMD | 0.0116 | 0.8438 |
| **MN** | SLE | -0.001 | 1.0 |
| **MN** | TBMD | 0.0017 | 1.0 |
| **LN** | TBMD | 0.0027 | 0.9998 |

Supplemental Table 2 - **Comparison of FP area mode between diagnoses.** The upper part of the table shows the mean mode for FP area for each diagnosis group as well as the coefficient of variance (CV%) within each diagnosis group. The mode was calculated for each patient using kernel density estimation and presented as the mean for each diagnosis group. The lower part shows a statistical comparison of the mean mode of FP area between the diagnosis groups. Normality was tested by Shapiro-Wilk and all data passed the normality test. ANOVA and Tukey's HSD was used to compare the group with adjusted p-values for multiple testing. Positive p-values are denoted with "*".
